# The impact of containment measures and air temperature on mitigating COVID-19 transmission: non-classical SEIR modeling and analysis

**DOI:** 10.1101/2020.05.12.20099267

**Authors:** Di Liu, Qidong Tai, Yaping Wang, Miao Pu, Sikai Ge, Tingting Ji, Lei Zhang, Bo Su

## Abstract

Early non-pharmaceutical interventions (NPIs) are crucial to prevent and control of COVID-19 pandemic. We established a stochastic non-classical SEIR NPIs model (ScEIQRsh) which can quantify the three kinds of NPIs measures simultaneously to mimic the clustered intra-family or intra-acquaintance spreading pattern of COVID-19 under the effective integrated NPIs in Mainland China. Model simulation demonstrated that measures to diminish contactable susceptible (Sc), such as home confinement, travel constraint, social distancing etc. and measures to avoid delay of diagnosis and hospitalized isolation (η) were more effective but consumptive than contact tracing (κ, ρ). From fitted model by MCMC method, the proportion of asymptomatic infectors was 14.88% (IQR 8.17%, 25.37%). The association between air temperature and the fitted transmission rate (β) of COVID-19 suggests that COVID-19 pandemic would be seasonal with the optimal temperature range of 5°C-14°C and peak of 10°C for spreading, and vaccine is indispensable to ultimate prevention COVID-19.

## Introduction

COVID-19 is spreading worldwide since January 2020 and has become a global health threat. World Health Organization (WHO) has designated this novel coronavirus as SARS-COV-2(*1, 2*) and declared it as a pandemic on March 11, 2020(3). As of May 19, 2020, there are a total of 4781963 accumulative confirmed cases in 214 countries and 84503 accumulative confirmed cases in China. During the spreading of COVID-19 in Mainland China, integrated social interventions including lockdown of high epidemic areas, travel restriction, social distancing, contact tracing for quarantine, and hospitalized isolation were undertaken to prevent the COVID-19 spreading in all the provinces after the lockdown of Hubei province on the Jan 23, 2020 in emergency (*4*). The effectiveness of NPIs has been proved in some infectious diseases, like influenza(*5-7*) and SARS(8). The NPIs are vitally important especially for COVID-19, because of the so fast spreading and the subsequent insufficiency of health-care resources leading to high mortality(*9*). Almost all the provinces were in semi-closure and executed with community closed management, meanwhile multiple provincial contact tracing groups were established to notify the close contacts to be in quarantine at home or indicated shelters. And for the suspected and diagnosed COVID-19 patients, early detection, early reporting, early isolation and early treatment were adopted according to the Chinese Clinical Guidance for COVID-19 Pneumonia Diagnoses and Treatment version 6(*10*).

Under the effective social NPIs, the spreading pattern of COVID-19 in most provinces of China was characterized of clustered or intra-family transmission(*11*) and mitigation by NPIs(*12*). To simulate the early spreading and early social intervention, we established a novel non-classical SEIR NPIs model, named ScEIQRsh (contactable Susceptible-Exposed-Infected-Quarantined-Removed) model to depict COVID-19 spreading. The classical SIR or SEIR (Susceptible-Exposed-Infected-Removed) epidemic model assumes that infectors mix up with all the susceptible per day, whereas ScEIQRsh model assumes that infectors mix up with the contactable susceptible per day. The contactable susceptible indicates the family members, relatives, co-workers, friends and some contactable strangers for obtaining daily necessities of the infectors. Several studies assessed one single aspect of NPIs, such as travel restriction(*13, 14*), airport screening(15, *16*), social distancing(*17*), and contact tracing(*18*) by classical SEIR based model without consideration of the other NPIs’ influence on the transmission property of COVID-19. We aim to estimate the effectiveness of integrated NPIs to simulate COVID-19 restricted spreading.

With the NPIs taken into account, the other clinical features, such as incubation period, the communicable period and the proportion of so-called asymptomatic infectors of COVID-19 were evaluated by this comprehensive model. The meteorological impact on COVID-19 transmission was still controversial(*19, 20*), and with this new ScEIQRsh model, we depicted the association of air temperature with COVID-19 transmission rate (β) among the provinces of China.

## Results

### Fitting the COVID-19 transmission with integrated social NPIs

The ScEIQRsh model can be well fitted with both the reported number of daily accumulative confirmed cases and close contacts being in quarantine in 29 provinces of Mainland China (Fig. 2, Fig. S1). The predicted daily Rh and Q compartments were coincident with the provincial reported numbers. From the fitting curves, the accumulative confirmed cases reached the plateau and stop increasing within 25-30 days, which indicated that the COVID-19 transmission was mitigated after emergency response to COVID-19 in all the provinces. The median transmission rate (β) for 29 provinces were 10.22 (IQR 8.47, 12.35), which implied about 10.22 person would be infected by one infector in case that the susceptible are mostly acquaintances and Sc is extrapolated to infinite (Fig. 3A, Table S2-S3). The index for overall assessment of NPIs effectiveness, Rvq, was smaller than one in every province with a median of 0.64 (IQR 0.62, 0.66), which demonstrated that implemented NPIs was intensive enough to make the number of free infectors decrease (Fig. 3B). The smallest Rvq (0.53±0.02) and largest Rvq (0.75±0.02) was observed in Shandong and Hubei province (Wuhan locates on), respectively, which implied the difficulty of NPIs with greater burden of contact tracing if the spreading has expanded with too many infectors in Hubei.

**Fig. 1.**
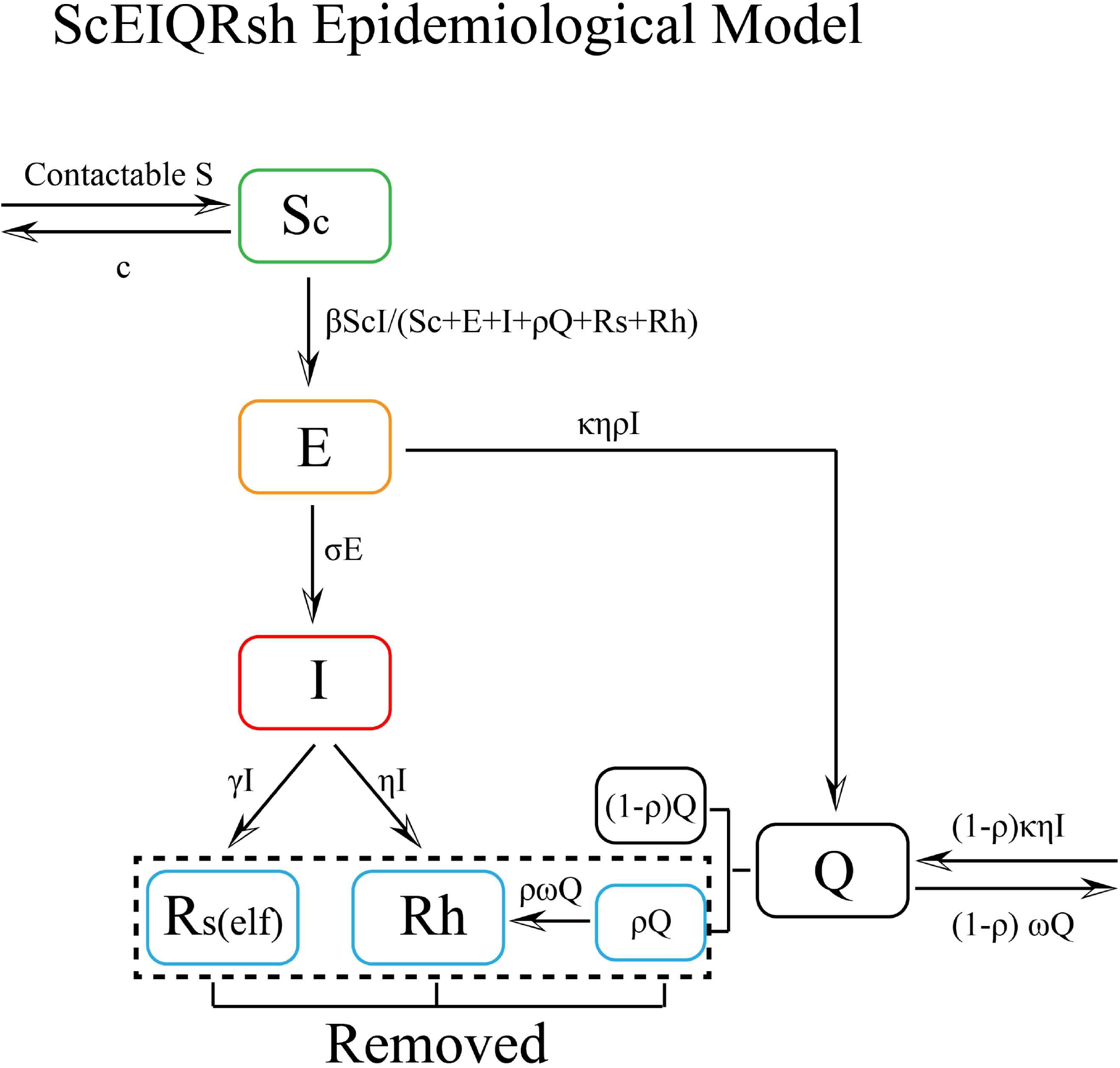
Flow diagram of ScEIQRsh epidemiological model. Six compartments: contactable susceptible (Sc), exposed individuals (E), infected individuals who were outsides of the public health measures (I), close contacts being in quarantine (Q), self-recovery individuals (Rs) and the cumulative hospitalized individuals (Rh). The flow velocities between the compartments are indicated.

**Fig. 2.**
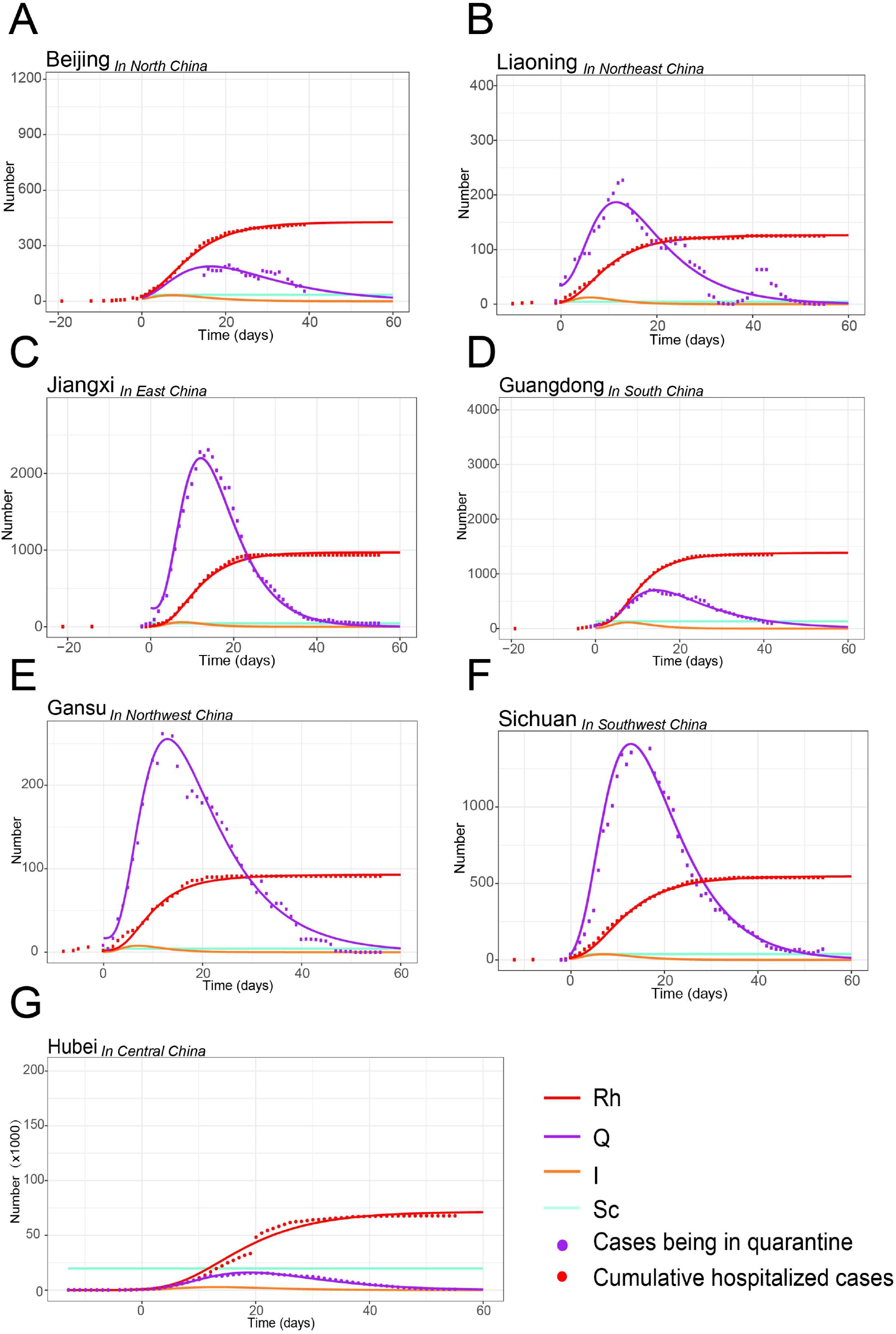
A-G: The representative fitting curves of both the number of daily cumulative confirmed cases and close contacts being in quarantine in provinces of Mainland China (Day 0, the 23^rd^, Jan, 2020).

**Fig. 3.**
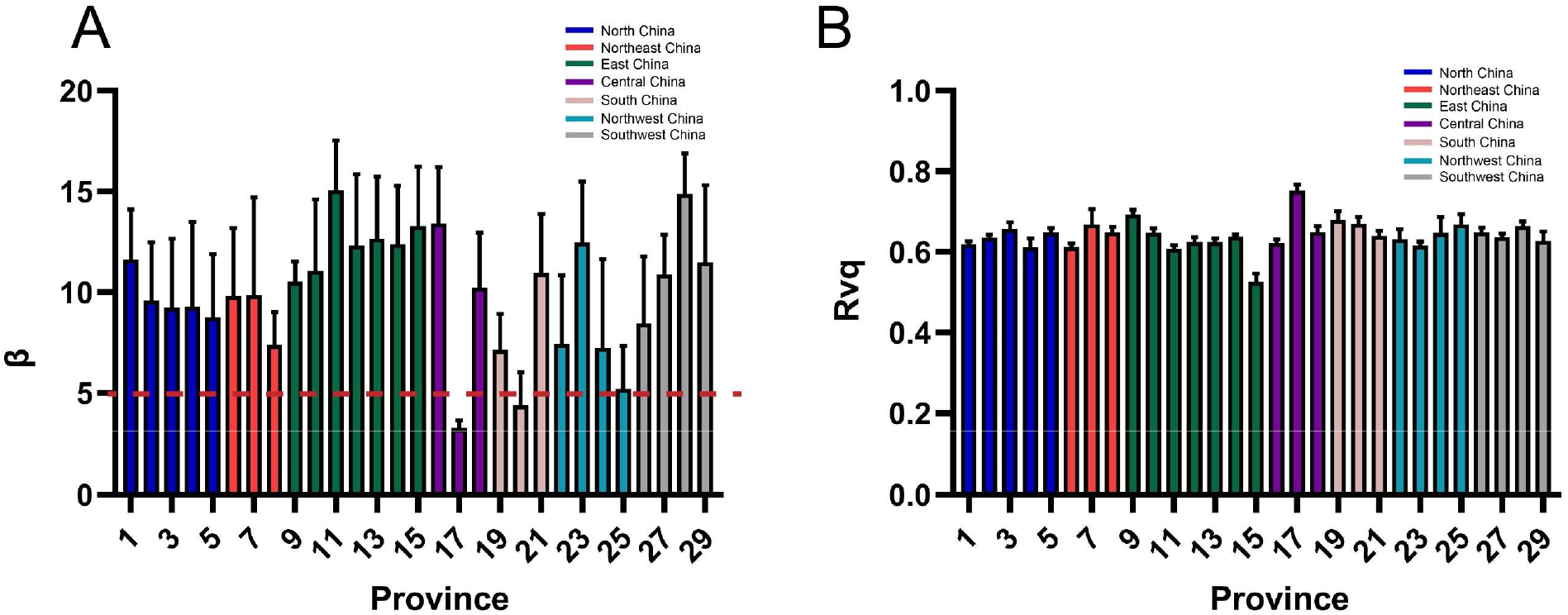
The transmission rates of COVID-19 and Rvq for 29 provinces in the fitted ScEIQRsh model. In figure A and B, the number on the X axis represents the provinces of each geographical region. North China: 1-5; Northeast China: 6-8; East China: 9-15; Central China: 16-18; South China: 19-21; Northwest China: 22-25; Southwest: 26-29. A. The transmission rates of COVID-19 among acquaintances for the 29 provinces grouped by geographical regions. B. The mean value of Rvq in the 29 provinces. If Rvq<1, the intensity of non-pharmaceutical-intervention is enough to stop propagating of infectors, the infectors will decrease.

### Assessment of NPI measures mitigating the spreading of COVID-19 and suppositional simulation

Three independent parameters, contact rate (Sc(c)), η and surveyQ, were crucial for stopping the spreading chain and evaluated separately for NPIs effectiveness. Based on our model, the median Sc was estimated as 26.98 (IQR:13.97, 54.57) with the highest in Hubei and lowest in Neimenggu province (Table S3). The median Sc(c) for 29 provinces was accordingly 6.84E-07 (IQR 3.77E-07, 1.44E-06) (Fig. 4A). The κ and ρ can be used to assess the effectiveness of CCT, and the median κ of 29 provinces was 42.0 (IQR 27.83, 60.78), suggesting that 42 close contacts of one infector had been excavated averagely by CCT groups (Fig. S2A). The COVID-19 positive rate (ρ) in close contacts was speculated as 0.98% (IQR 0.47%-1.60%), ranged from 0.03%-5.10% (Fig. S2B), which was quite close to the WHO-China joint report of 0.9%-5% in China(*12*). SurveyQ, the product of κ times ρ, was 0.39 (IQR 0.22, 0.55), which indicated that 0.39 positive cases were found in close contacts of CCT averagely for each confirmed infector (Fig. 4C). The median for velocity of hospitalized isolation for infectors (η) was 0.69 (IQR 0.47, 0.87) among 29 provinces, with η value greater than 0.85 in Jilin, Heilongjiang, Shanghai, Jiangsu, Hubei, Hunan, Guangxi and Guizhou province (Fig. 4B).

To illustrate the influence of NPI measures on COVID-19 transmission for ScEIQRsh model, we arbitrarily adjusted Sc (c), κ, ρ, and η value with representative 30% or 50% up/down-regulation to simulate the suppositional spreading situation. If Sc (c) were 30% or 50% enlarged or η reduced, the eventual accumulative hospitalized cases (Rh) would strongly increase, the peak of infectors (I) would be brought backward and vice versa (Fig. 4D-4G). In case of enlarged κ or ρ, the eventual accumulative number of confirmed COVID-19 would diminish and the plateau of Rh would be brought forward (Fig. 4H-4I, Fig. S2C-S2F). On the same adjusted amount, the effectiveness of Sc (c) and η on spreading prevention were stronger than that of CCT parameters, κ and ρ.

**Fig. 4.**
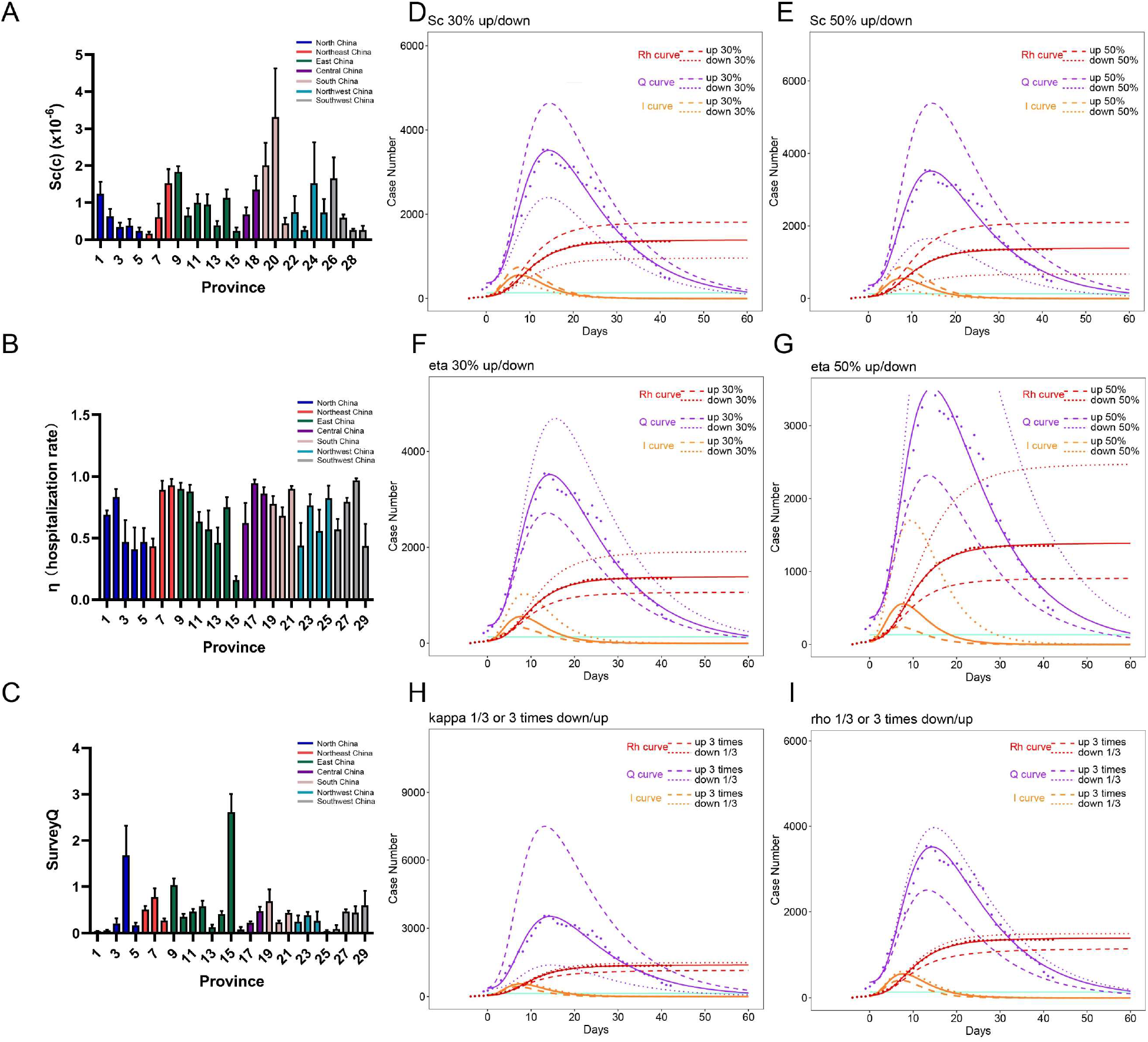
Evaluation of effectiveness for the 3 categories of NPI measures to mitigate COVID-19 spreading and suppositional simulation. A: The contact rates (Sc/N) were smaller than 4*10^-6^ in 28 provinces except Hubei. D-E: The simulated Rh, Q and I curves were sketched when adjusted contact rate with 30% or 50% up/down-regulation. B: The hospitalization rate and pace, η, of the 29 provinces. F-G: The influence on Rh, Q and I curves after adjustment of η with 30% or 50% up/down-regulation. C: The quality of contact tracing, surveyQ was presented among 29 provinces. H-I: The influence on Rh, Q and I curves after adjustment of κ and ρ to 3 times or 1/3.

### Blind zone of contact tracing and asymptomatic infectors in NPIs strategy

With the integrated social NPIs, COVID-19 transmission mostly occurred between free infectors and acquaintances, as well as a few strangers whom the infectors have to contact for daily necessities in China. In a typical clustered intra-acquaintance transmission, the index case transmitted to four of his family members and one friend directly, and the friend’s family indirectly within half a month(*21*) (Fig. 5A). After emergency response, multiply provincial contact tracing group were established to investigate the contact history of each confirmed infectors and notified the close contacts to be in self-quarantine at home or in shelters. CCT can easily find close contacts of acquaintances, but be inefficient in finding transmission among strangers in public space. For example, a salesman index transmitted to two unacquainted salesmen in other sales areas sequentially without gathering in a large mall, and a customer without any inquiry and purchase was infected by one of the transmitted salesmen after 30-minutes lingering (Fig. 5B). This transmission chain in strangers could not be easily found by contact tracing, and was only revealed after all the participants appeared symptoms (Fig. 5C).

**Fig. 5.**
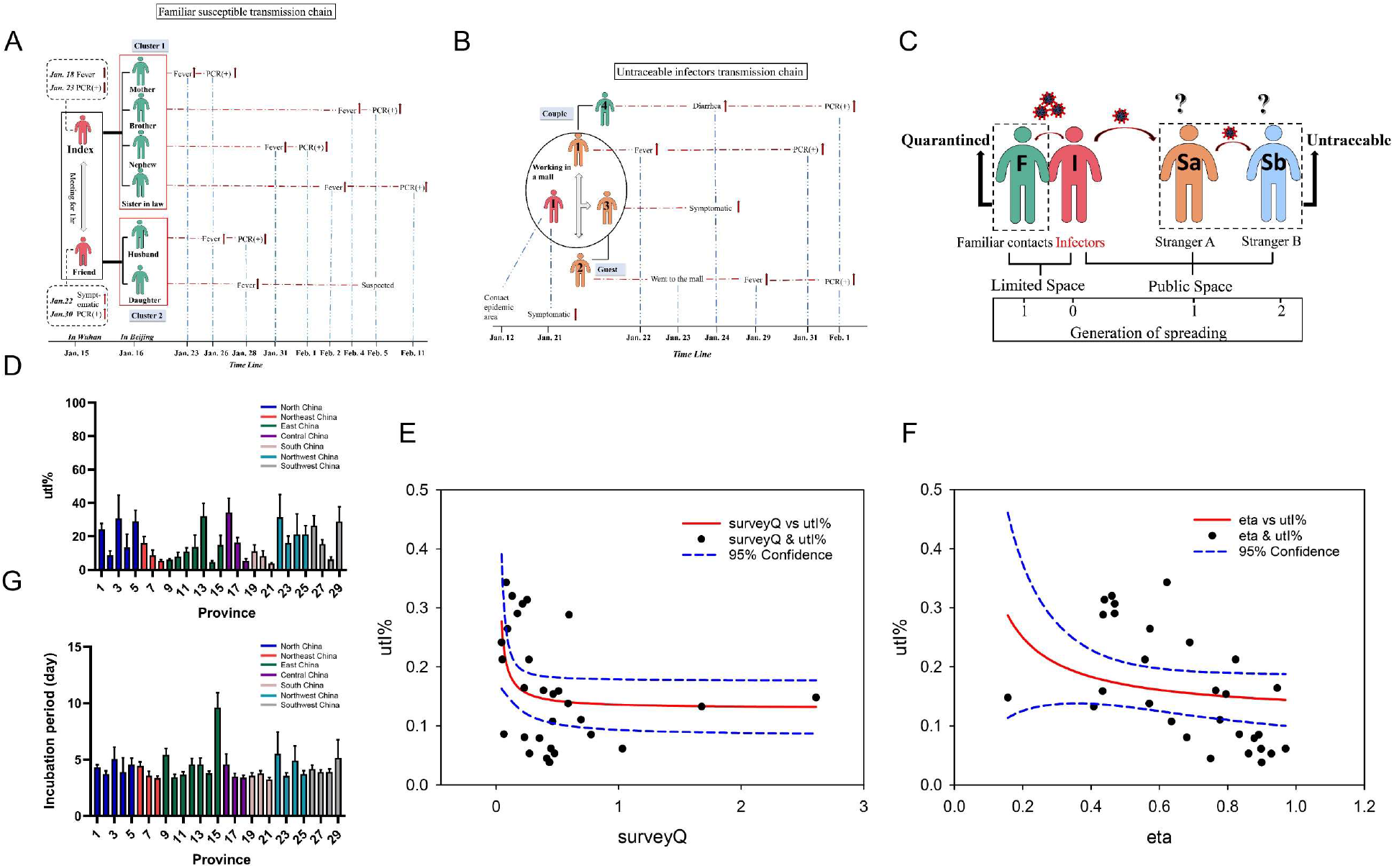
The transmission patterns of COVID-19 under social NPIs and the association of untraceable infectors with surveyQ/ η. A: A representative example of intra-family and intra-acquaintance transmission pattern of COVID-19 in Beijing. B: A representative example of COVID-19 transmission pattern among strangers in a large mall. C: Stranger-stranger transmission is the blind zone of contact tracing. D: The median proportion of asymptomatic infectors among 29 provinces. E-F: The nonlinear association between the ration of untraceable infectors and survey / η. Higher η or surveyQ could reduce the ration of untraceable infectors. But as the surveyQ increasing, the proportion of untraceable infectors would be constant. G: The median incubation period of COVID-19 among 29 provinces.

Another blind spot of contact tracing is about asymptomatic infection. Rs compartment in the ScEIQRsh model consists of the self-recovery individuals who have never be hospitalized and not registered in healthcare system mostly because of asymptomatic infection or mild symptoms. The calculated proportion of asymptomatic and mild-symptom infectors without hospitalization (untraceable infectors, utI%) by ScEIQRsh model was 14.88% (IQR 8.17%, 25.37%), ranged from 3.92%-34.36% across 29 provinces, which implied that average 14.88% COVID-19 patients couldn’t be found out with social NPIs, and the average proportion of asymptomatic in COVID-19 patients should be smaller than 14.88% (IQR 8.17%, 25.37%) (Fig. 5D). The higher surveyQ of CCT can only reduce but not eliminate utI% (Fig. 5E), but high η could decline the utI% constantly (Fig. 5F). Hence, contact tracing is not sufficient to find all the infectors, especially in stranger-stranger transmission and asymptomatic infection. An integrated NPI strategy is strongly recommended; as we earlier simulated, Sc (c) and η have stronger prevent effectiveness, which means staying home to diminish contacts, making diagnosis and hospitalized isolation as soon as possible.

### The incubation period and communicable period of COVID-19 in ScEIQRsh model

Computed with the fitted model, the median incubation period was calculated with sigma and eta (incubation period ≈1/α+1/η), of 4.16 days (IQR 3.60, 4.71) (Fig. 5G). The communicable period of COVID-19 was infectious period related to asymptomatic and some mild-symptom infectors, with a median of 6.77 days (IQR 4.53,10.36) calculated as 1/γ (Table S2). The median time from individuals exposed to SARS-COV-2 to being contagious was 2.39 days (IQR 2.26, 2.56) determined by 1/σ (Table S2).

**Fig. 6.**
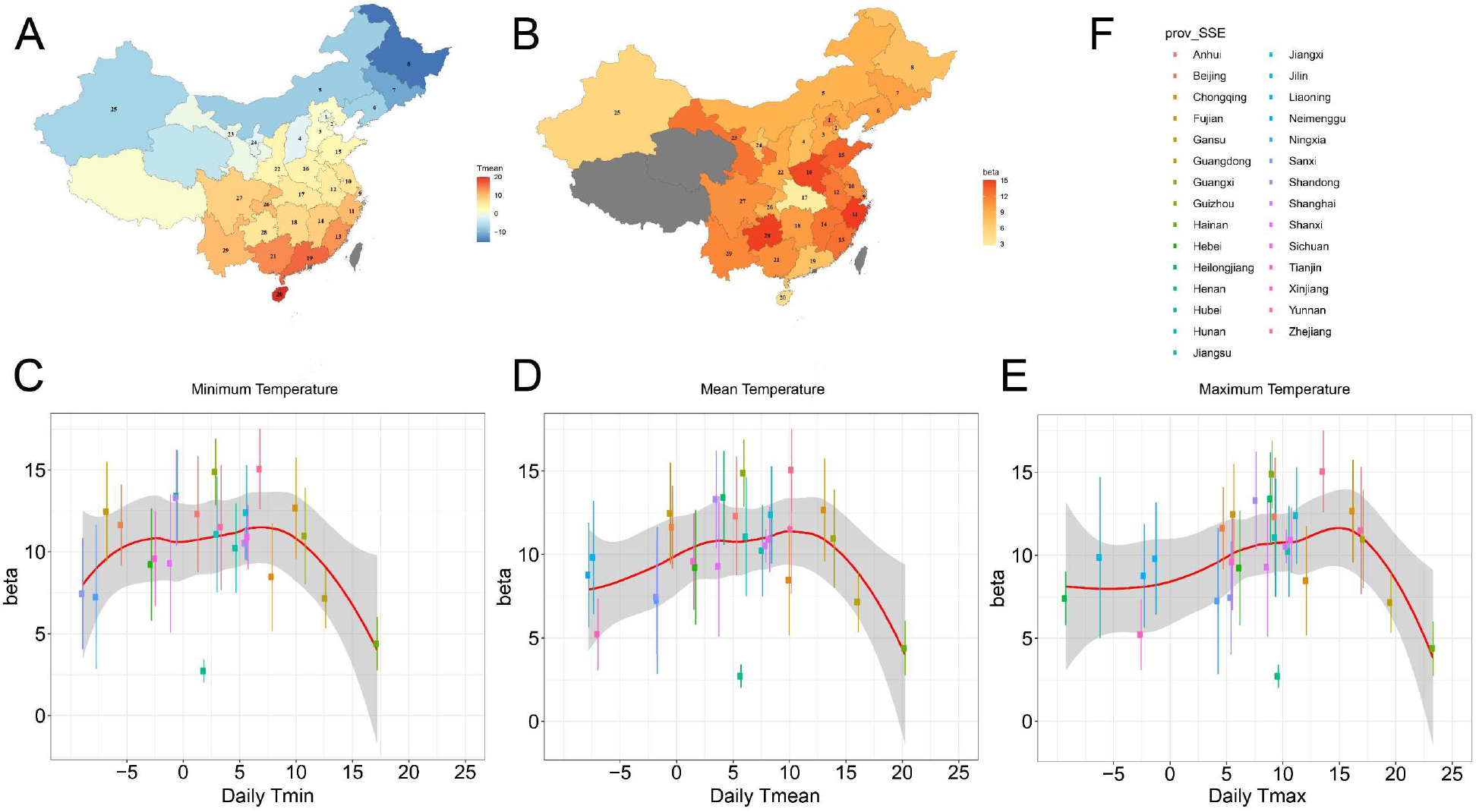
The impact of minimum, mean and maximum air temperature on the transmission of COVID-19 in mainland China. A: Mapping the daily mean temperature from the 15^th^, Jan, 2020 to the 15^th^, Feb, 2020 in 29 provinces of Mainland China. B: Mapping the transmission rate of COVID-19 in 29 provinces of Mainland China. C, D, E: The association between daily minimum (C), mean (D) and maximum (E) temperature and the transmission rate, β of COVID-19 depicted by LOESS, sharing similar shape of curves. F: The legend for figure C-E. One dot represents the minimum, mean or maximum temperature of one province.

### The influence of air temperature on the transmissibility of SARS-COV-2

The span of mean air temperature for every province was from -15 °C (5 °F) to 20.25 °C (68.45 °F) during the COVID-19 spreading period of Jan to Feb 2020 in China (Fig. 6A-6B). The nonlinear association between transmission rate β and air temperature was depicted by LOESS fitting. As the daily air temperature increased from subzero (0°C, 32°F), the value of β raised gradually until the air temperature went up to a peak of 7°C(44.6°F) for minimum daily temperature, or 10°C (50°F) for mean or 15°C (59°F) for maximum daily temperature respectively (Fig. 6C-6E), and then declined sharply as the temperature continued to raise. We observed transmission rate β was higher than 11 in the range of 5°C-14°C (41°F-57.2°F) for mean air temperature, which may be most suitable for COVID-19 spreading.

## Discussion

With the experience of SARS in 2002, the first-level public health emergency response for COVID-19 was launched on the Jan 23, 2020 in Mainland China. The integrated NPIs are as follows(4): 1) lockdown of high epidemic areas; 2) extension of the Spring Festival to the Feb 2, 2020; 3) inter- and intro-province travel restraint; 4) closing public places, avoiding gatherings and staying home; 5) environmental disinfection and personal protective measures including wearing masks; 6) social distancing; 7) setting up provincial contact tracing groups and contact tracing for each confirmed case or imported infector; 8) notification of close contacts to be in quarantine for 14 days at home or shelters; 9) identification of suspected COVID-19 in cough and fever clinics separately; 10) hospitalized isolation for every confirmed case; 11) preventing healthcare-associated infections. Among them, 1-6 diminished Sc or c, 7-8 contributed to κ and ρ, 9-11 contributed to η in the ScEIQRsh model.

The classical SEIR based NPI model usually have to be staged to simulate the propagating stage and NPI stage separately, instead, ScEIQRsh model can well fit the realistic provincial epidemic and NPI data of COVID-19 in China entirely without halfway adjustment of the parameters. The assumption of ScEIQRsh model makes it a model to depict intra-acquaintance spreading pattern, and if Sc were replaced with all the susceptible, it could degenerate into classical SEIR model. A report of the “WHO-China Joint Mission on COVID-19 ” claimed that human-to-human transmission of COVID-19 largely occurred intra-family in China(*12*). In Guangdong and Sichuan province, about 78%-85% of clustered infections were within families(*12*). And in Beijing, there are 176 clustered cases among 262 confirmed cases and 133 (50.8%) were clustered intra-family cases(*11*). The implementation of effective social NPIs would change the spreading pattern of COVID-19 into intra-acquaintance spreading, in which acquaintance refers to the family members, relatives, friends and co-workers, as well as some contactable strangers for obtaining daily necessities. That’s the reason why ScEIQRsh model can well fit both COVID-19 epidemic and NPI data of China.

According to the ScEIQRsh model, Sc (c) and η are more effective to diminish the eventual accumulative number of COVID-19 cases than CCT parameters κ and ρ. We can summarize the goal of measures 1-6 as “staying home and going out with precautions” for the susceptible, and measures 9-11 as “quickening diagnosis and hospitalized isolation” for infectors. In case of insufficiency of medical resources, the better way to improve η could be enlargement of the laboratory capacity for SARS-COV-2 testing or building makeshift hospitals to increase bed capacity (*22*). Contact tracing is also helpful for mitigating COVID-19 spreading, which can be regarded as a reminder to be more precautious for close contacts (quarantine for targeted susceptible) than the common susceptible. The surveyQ (κ•ρ) of CCT could be improved with adding more CCT staffs, loosening the criteria of close contacts in CCT, broadening SARS-COV-2 testing to close contacts, or using digital tools. Nevertheless, lockdown and stay-at-home affects the society and economy seriously, contact tracing is an alternative option with less quarantine and consumptions.

Additionally, the asymptomatic infectors with contagiousness are source for recurrence of COVID-19 epidemic. We demonstrated that the median and highest proportion of asymptomatic infectors were 14.88% and 34.36% respectively. It was consistent with the reported 18% in 700 infectors never showed symptoms on *Diamond Princess* by Kenji Mizumoto *et al*(*23*) and claim of 30.8% asymptomatic cases in 565 Japanese citizens evacuated from Wuhan by Hiroshi Nishiura *et al* (24). The calculated median incubation period of COVID-19 was 4.16 days (IQR 3.60, 4.71), which was close to the reported 3 days in a study of including 1,099 laboratory-confirmed cases by Zhong N.S *et al* (25) and 5.1 days in 181 confirmed cases of COVID-19 by Lessler Justin *et al* (*26*). The median communicable period (1/γ) of asymptomatic infectors was computed as 6.77 days (IQR 4.53, 10.36), which was near the 9.6 days in 24 asymptomatic infectors reported by Shen H.B *et.al*(*27*).

Previous studies about meteorological impact on the SARS-COV-2 transmissibility were still controversial. Wang Mao *et al*(*19*) and Sajadi MM *et al*(*20*) reported that regions in 30-50° N’ corridor with 5°C-11°C (41°F-51.8°F) average temperature had increased COVID-19 transmission, but Canelle Poirier *et al*(*28*) indicated no significant correlation between high temperature/humility and COVID-19 transmission. We found the transmission rate β increased as the minimum, mean, maximum temperature rose from -5°C (23°F) and reach the peak at 7°C (44.6°F), 10°C (50°F), and 15°C (59°F), respectively, and then started to decline at higher temperature across the 29 provinces. It is coincident with the curves reported by Wang Mao *et al*(*19*), who have claimed that the peak of accumulative cases of 492 cities appeared at the minimum, average and maximum temperature of 6.7°C(44.06°F), 8.72°C(47.7°F) and 12.42°C(54.36°F) respectively. The optimal mean temperature ranges for COVID-19 transmission was 5°C-14°C (41°F-57.2°F). Lowen Ac *et.al* proved that influenza virus transmission by droplets was greater and the peak duration of virus shedding lasted longer at 5°C (41°F) than 20°C (68°F)(*29*). Indeed, we observed the β of COVID-19 at 5°C (7.39±1.62) was higher than β at 20.25°C (4.39±1.63). Another similar infectious disease, SARS was found with higher transmission in temperature<24.6°C than >24.6°C circumstance in Hong Kong(30). From the curve we illustrated, we could suspect the transmission rate would further reduce at higher temperature over 20.25°C (68.45°F). It can be expected that COVID-19 pandemic spreading would mitigate for the northern hemispheres in the coming summer, but it is very likely to become a seasonal infectious disease. The broader range of air temperature for optimal COVID-19 transmission strongly suggests the necessity and urgency of vaccine at present or in the future.

This study only included the epidemic date of Mainland China for model validation because we cannot access the NPIs data, e.g. close contacts of other countries. This model could be fitted even if the NPIs data is sparse, but with less accuracy. The limited latitude spans made the range of air temperature not so broad, especially for higher temperature, though the association of air temperature with COVID-19 transmission rate was informative and suggestive. The proportion of asymptomatic infection was smaller than some other reports, which may due to the meaning of asymptomatic infectors has changed. The asymptomatic infectors mentioned in latter reports usually indicated SARS-COV-2 RNA positive individuals without symptoms, and this could happen in the early infection phase of SARS-COV-2.

In conclusion, we provided a new tool for quantitative assessment or prediction of the effectiveness of NPIs strategy, which was validated with both spreading and social intervention data of COVID-19 in China. The effectiveness of the three kinds of NPI measures in the fitted model and the influence of air temperature on COVID-19 transmissibility were quantitatively assessed, which provided new evidence to decision making of NPIs strategy for COVID-19 prevention and control. This model is also applicable for any other regions because the proportion of acquaintances and strangers could auto-adjust by the fitting process, and it is also suitable for other similar infectious disease.

## Materials and Methods

### Development of the dynamical non-classical SEIR model for COVID-19

The developed comprehensive ScEIQRsh model contains six compartments named as Sc (contactable susceptible), E (the exposed to SARS-COV-2), Q (daily close contacts being in quarantine), I (infectors outside of the healthcare system), Rh (accumulative hospitalized infectors), and Rs (self-recovery individuals with asymptomatic infection or mild symptoms who have never be hospitalized and registered in healthcare system) (Fig. 1). Sc represents the contactable susceptible under the social NPIs, such as lockdown, social distancing, cancelling gatherings, closing public places, which is set as a random variable in the model. Q reflects the contact tracing (CCT) for quarantine, and Rh reflects the isolation of confirmed and hospitalized infectors. E, I, and Rs compartments were outside of the healthcare system. The model can be mathematically described as five differential equations (details in the supplementary methods).

### Parameters in ScEIQRsh model

The definition and initial range of model parameters of β, σ, γ, κ, ρ, ω, η and Sc in the model were listed in Table S1. β was the transmission rate dependent of the property of SARS-COV-2. σ and γ were associated with the intrinsic incubation period and communicable period of COVID-19. κ, ρ and ω were associated with contact tracing and quarantine. η reflected the pace of confirmed diagnosis and hospitalized isolation for infectors. Other indexes could be calculated from the solved model, such as the contact rate (Sc(c)), the quality of CCT (surveyQ), the proportion of untraceable infectors (approximately equate to the asymptomatic) (utI%), the incubation period and communicable period of SARS-COV-2, and among them, Sc(c) reflects the proportion of Sc in population under the integrated NPIs, surveyQ represents the quality of contact tracing. Rvq is defined as the ratio of transmission velocity to remove velocity when Q reached the apex, which is used to evaluate the overall effectiveness of integrated NPIs.

### Data and model validation

The daily accumulative confirmed cases and daily close contacts being in quarantine of COVID-19 from January to March, 2020 through the 31 Provincial Health Commission of Mainland China were collected. Xizang and Qinghai province were excluded because of few cases, with only one and 18 confirmed cases respectively. Almost all the diagnosed cases were hospitalized in isolation wards simultaneously according the Guidance, thus the reported confirmed cases were just the hospitalized infectors in China. Details of the criteria of confirmed cases and close contacts in quarantine were clarified in the supplementary methods. We fitted both the reported accumulative confirmed cases and daily close contacts in quarantine with Rh and Q compartments by ScEIQRsh model for each province. The value of parameters was random sampled with one of Markov Chain Monte Carlo (MCMC) method, Metropolis-Hastings (M-H) algorithm, and documented under an appropriate tolerance of best fitting with at least 100000 iterations of 0.1 step size from 0 to 60 days with burn-in of 50000 iterations for every province of Mainland China.

### Air temperature of provinces in China during the COVID-19 spreading

The minimum, mean, maximum air temperature of each province was collected from the National Meteorological Administration from Jan 15, 2020 to Feb 15, 2020 (one week before and three weeks later of Jan 23, 2020 (Day 0)). The LOESS regression was used to depict the relationship between the air temperature of 29 provinces and COVID-19 transmission rate in ScEIQRsh model.

### Statistical analysis

The enrolled 29 provinces for model validation were separated into seven geographical regions, named as North, Northeast, East, Central, Northwest, South and Southwest China. Both curves of Rh and Q was simultaneously fitted with the raw data by least squares method with tolerance of 1.05, and the cost function was SSE/SST (Sum of Squares for Error/Sum of Squares for Total). The proposal distribution for accept-reject in M-H algorithm is a Bernoulli distribution from the comparison of cost function of curve fitting in iteration. The data process was performed on R (version 3.6.1), and the “deSolve” R package was used as the solver of differential equations. The R source code can be found in *GitHub*.The parameters were denoted as mean±SD for each province, and the median (interquartile range, IQR: 25%, 75%) was used for describing across provinces.

## Data Availability

No additional data avaiable.

## Author contributions

DL and QT are joint first author, contributed equally to this article for drafting the manuscript. BS and LZ conceived and designed the study. QT, MP and YW collected the epidemiological data of each province in Mainland China. BS and DL analyzed the date with the help of SG, TJ, YW and MP. DL and QT drafted the manuscript. BS and LZ revised the manuscript critically. All authors approved the final manuscript. LZ and BS are co-corresponding author and the guarantors. The corresponding authors attests that all listed authors meet authorship criteria and that no others meeting the criteria have been omitted.

## Data and materials availability

The data are available on request.

Figures

## Supplementary figures

**Fig. S1.**
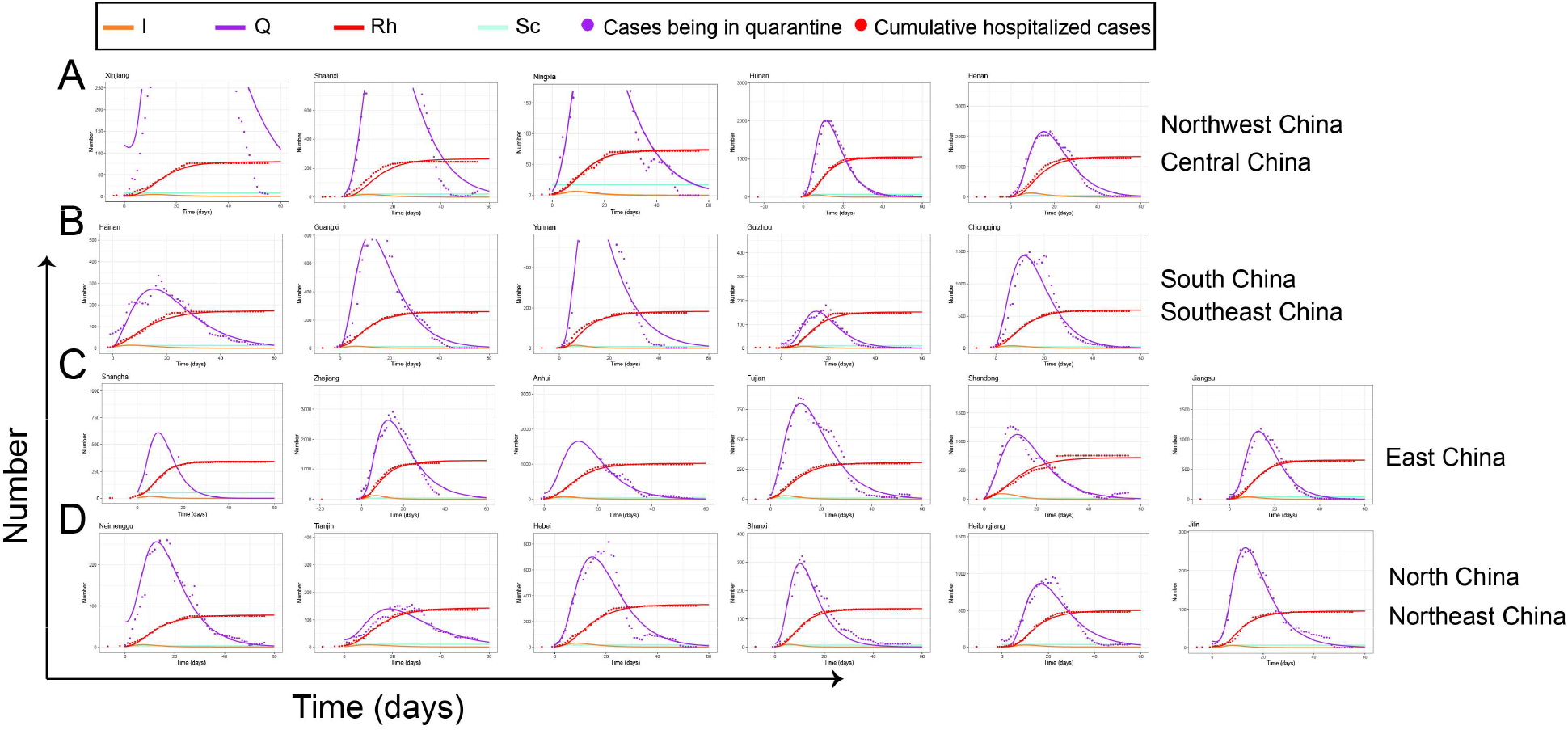
The fitting curves of both the number of daily cumulative confirmed cases and close contacts being in quarantine in 22 provinces of Mainland China (Day 0, the 23rd, Jan, 2020). A: The fitting curve of provinces in Northwest China-Xinjiang/ Shaanxi/ Ningxia and Central China-Hunan/ Henan. B: The fitting curve of provinces in South China-Hainan/ Guangxi, and Southwest China-Yunnan/ Guizhou/ Chongqing. C: The fitting curve of provinces in East China-Shanghai/ Zhejiang/ Anhui/ Fujian/ Shandong/ Jiangsu. D: The fitting curve of provinces in North China-Neimenggu/ Tianjin/ Hebei/ Shanxi and Northeast China-Heilongjiang/Jilin.

**Fig. S2.**
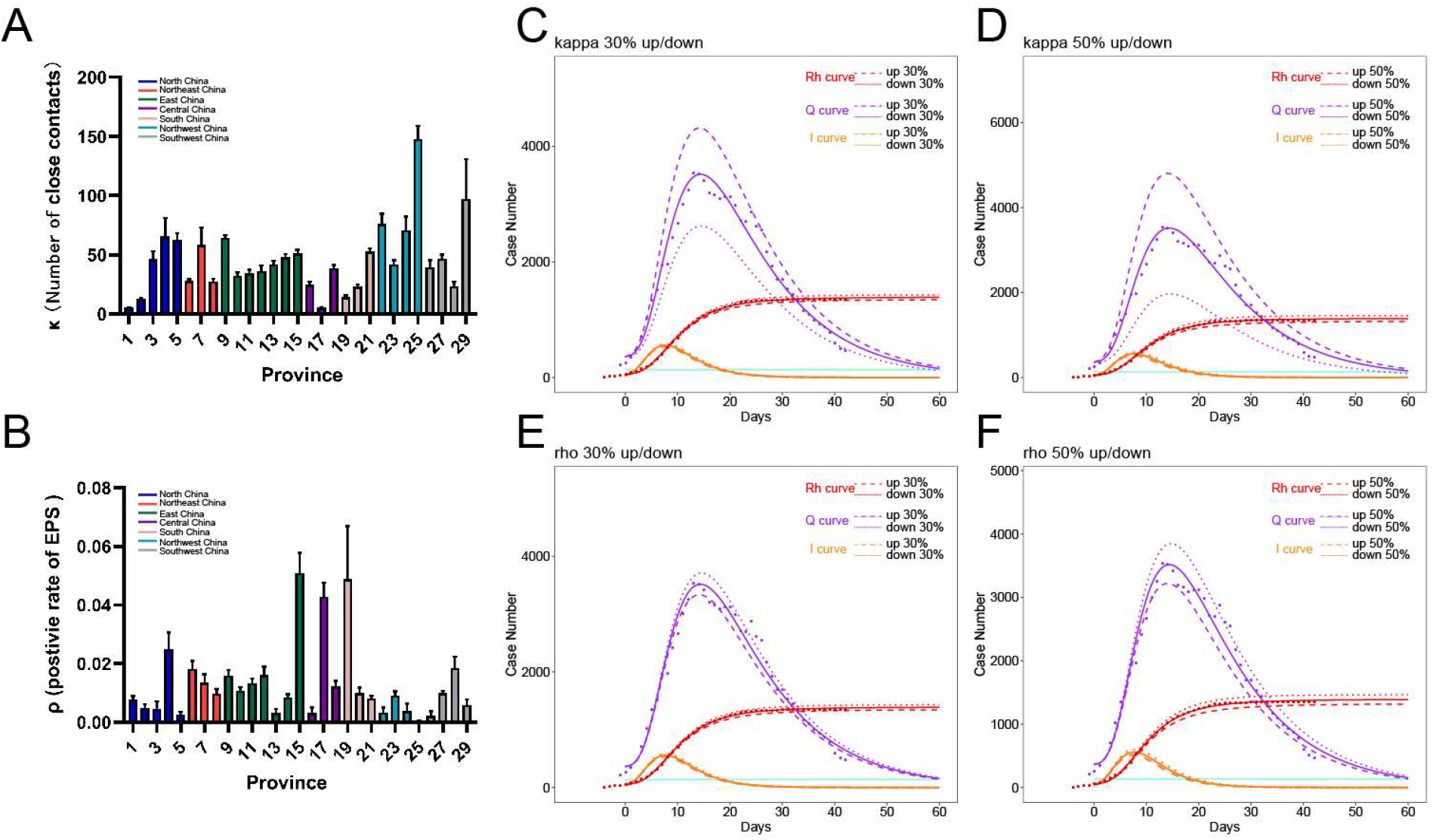
Suppositional simulation of contact tracing parameters, κ and ρ. A-B: The median κ and ρ was observed as different among 29 provinces. C-D: The influence on Rh, Q and I compartment after adjustment of κ by 30% or 50%. E-F: The simulated Rh, Q and I compartment after adjustment of ρ by 30% or 50%.

Supplementary Tables

**Table S1.** The definition and setting range of parameters in ScEIQRsh model.

| Parameters | Definition | Method | Setting range |
| --- | --- | --- | --- |
| $Sc$ | contactable susceptible under the social NPIs | MCMC | [1, 0.01N] |
| $\beta$ | transmission rate, the number of infected people by one infector | MCMC | [1,19] |
| $\sigma$ | transition rate from exposure to being contagious | MCMC | [0.27,0.5] |
| $\gamma$ | recovery rate of asymptomatic infector | MCMC | [0.04,0.3] |
| $\eta$ | hospitalization rate and pace of symptomatic infectors | MCMC | [0.001,0.999] |
| $\kappa$ | extent of epidemiological investigations | MCMC | [0,350] |
| $\rho$ | positive rate of COVID-19 in quarantined people | MCMC | [0,0.1] |
| $\omega$ | transited rate of quarantined people developing to contagious per day | MCMC | [0.07,0.6] |
| $Sc(c)$ | the proportion of contactable susceptible ($Sc$) under the interventive social prevention | $Sc/N$ | - |
| $Rvq$ | ratio of transmission velocity to remove velocity when Q reached the max | -* | - |
| $Ip_d$ | the time elapsed from exposure to SARS-COV-2 to the symptoms firstly apparent | $1/\sigma+1/\eta$ | - |
| utI% | the proportion of untraceable infectors, approximately equates to the asymptomatic | $\gamma/(\eta+\kappa\rho\eta+\gamma)$. | - |
| SurveyQ | the quality of contact tracing | $\kappa\rho$ | - |
| Cpd | the time for untraceable infectors with contagious among susceptible | $1/\gamma$ | - |

**Abbreviation:** Ipd: Incubation period; Cpd: communicable period; Asym%: Proportion of asymptomatic infectors; N: the total population of a province. \* Details in the supplementary methods

**Table S2.** The median value of parameters and indexes across 29 provinces of Mainland China by the fitted ScEIQRsh model.

| Variables | Median | IQR 25%, 75% | Range |
| --- | --- | --- | --- |
| $\beta$ | 10.22 | 8.47, 12.35 | 3.29-15.06 |
| $\sigma$ | 0.42 | 0.40, 0.44 | 0.33-0.48 |
| $\gamma$ | 0.15 | 0.10, 0.22 | 0.05-0.26 |
| $\eta$ | 0.69 | 0.47, 0.87 | 0.16-0.97 |
| $\kappa$ | 42.0 | 27.83, 60.78 | 5.35-147.79 |
| $\rho$ (%) | 0.9% | 0.4%-1.6% | 0.03%-5.10% |
| $\omega$ | 0.12 | 0.10, 0.15 | 0.07-0.21 |
| Sc | 26.98 | 13.97, 54.57 | 5.91-25525.54 |
| Sc(c) | 6.84E-07 | 3.77E-07, 1.44E-06 | 1.64E-07—4.33E-04 |
| Rvq | 0.64 | 0.62, 0.66 | 0.53-0.75 |
| Ipd | 4.17 | 3.60, 4.71 | 3.27-9.62 |
| utI% | 14.88% | 8.17%, 25.37% | 3.92%-34.36% |
| SurveyQ | 0.39 | 0.22, 0.55 | 0.04-2.61 |
| Cpd | 6.77 | 4.53, 10.36 | 3.91-19.90 |
| $1/\sigma$ | 2.39 | 2.26, 2.56 | 2.07-3.01 |

**Abbreviation:** Ipd: Incubation period; utI%: Proportion of asymptomatic infectors; Cpd: communicable period

**Table S3.**
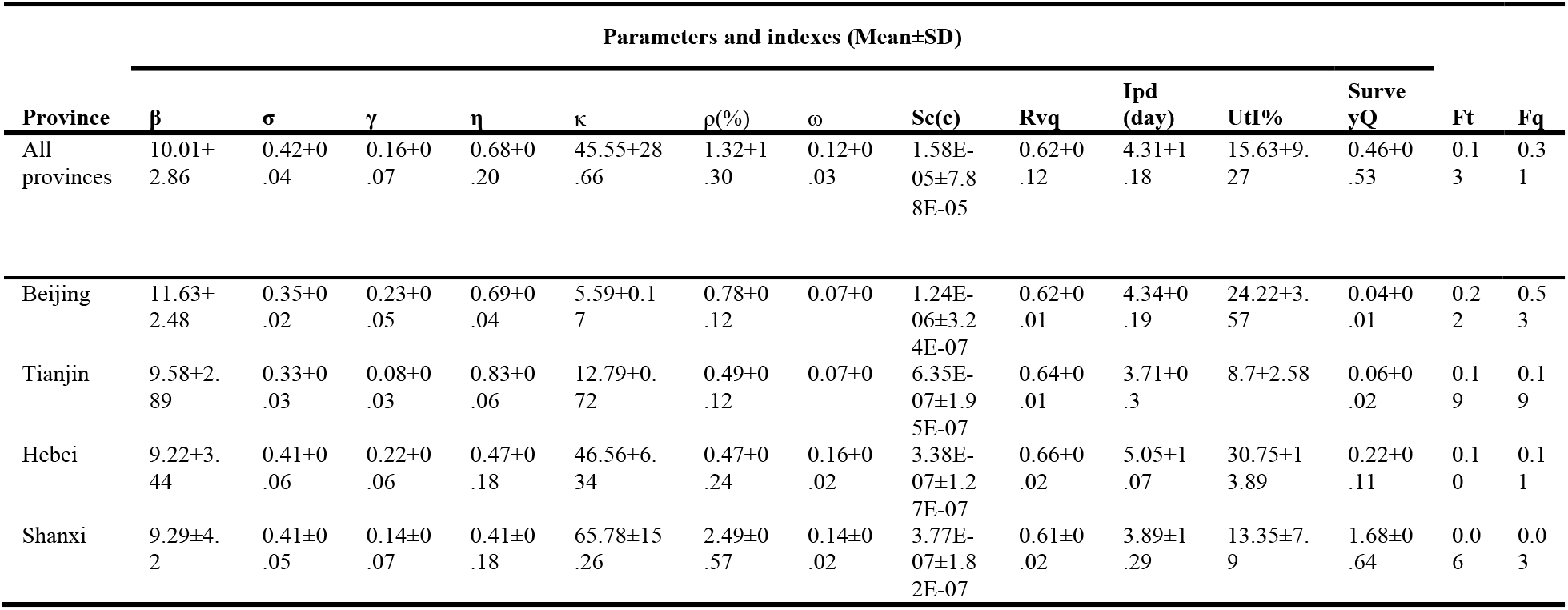
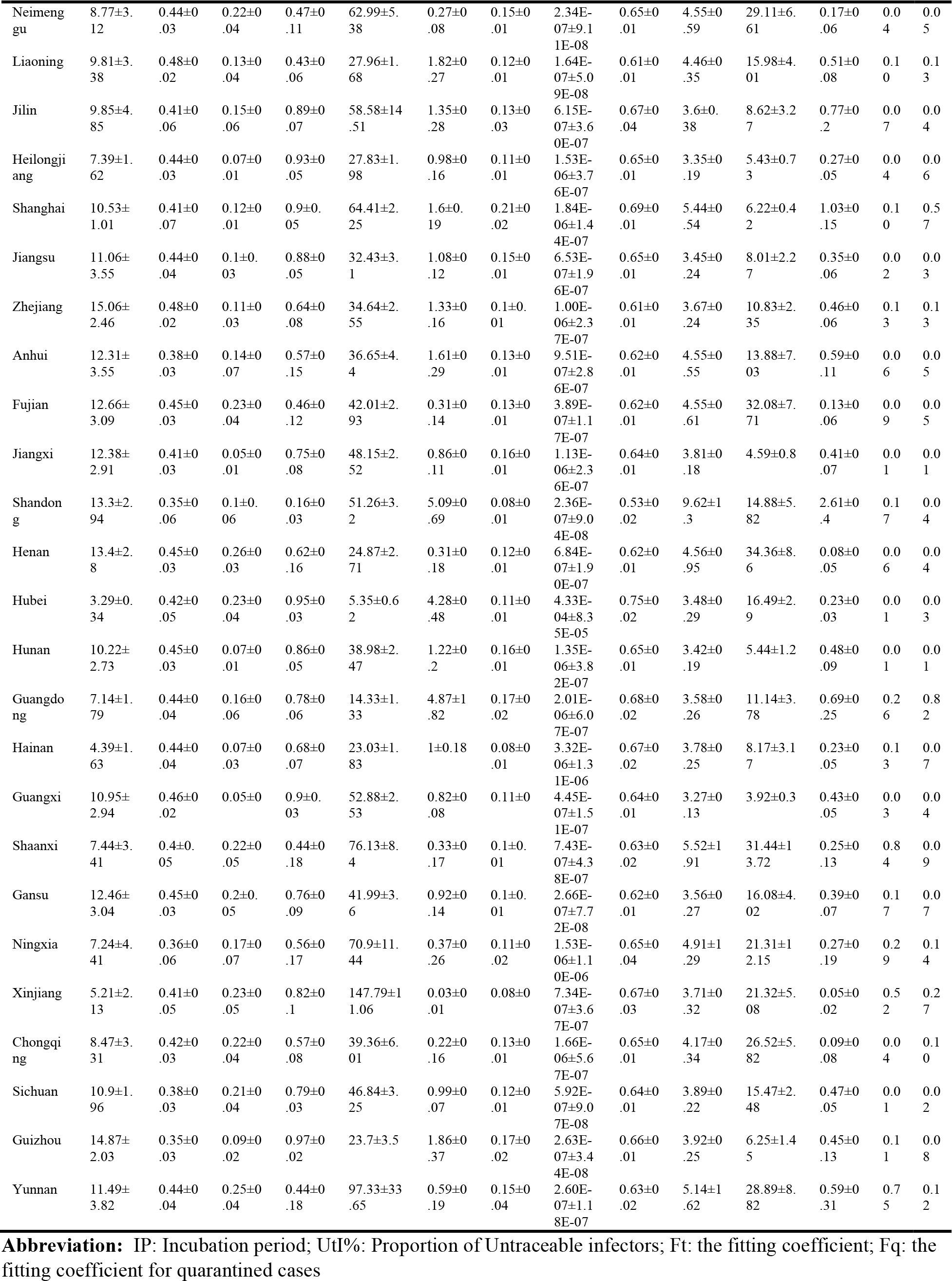
The mean value of parameters and indexes in each province of Mainland China.

## References

1. R. Lu, X. Zhao, J. Li, P. Niu, B. Yang, H. Wu, W. Wang, H. Song, B. Huang, N. Zhu, Y. Bi, X. Ma, F. Zhan, L. Wang, T. Hu, H. Zhou, Z. Hu, W. Zhou, L. Zhao, J. Chen, Y. Meng, J. Wang, Y. Lin, J. Yuan, Z. Xie, J. Ma, W. J. Liu, D. Wang, W. Xu, E. C. Holmes, G. F. Gao, G. Wu, W. Chen, W. Shi, W. Tans, Genomic characterisation and epidemiology of 2019 novel coronavirus: implications for virus origins and receptor binding. Lancet (London, England), (2020).

2. Wuhan Municipal Health Commission briefing on the pneumonia epidemic situation, 31 December 2019 (http://wjw.wuhan.gov.cn/front/web/showDetail/2019123108989).

3. WHO Director-General’s opening remarks at the media briefing on COVID-19 - 11 March 2020. (https://www.who.int/dg/speeches/detail/who-director-general-s-opening-remarks-at-the-media-briefing-on-covid-19-11-march-2020.)

4. W. Chen, Q. Wang, Y. Q. Li, H. L. Yu, Y. Y. Xia, M. L. Zhang, Y. Qin, T. Zhang, Z. B. Peng, R. C. Zhang, X. K. Yang, W. W. Yin, Z. J. An, D. Wu, Z. D. Yin, S. Li, Q. L. Chen, L. Z. Feng, Z. J. Li, Z. J. Fengs, [Early containment strategies and core measures for prevention and control of novel coronavirus pneumonia in China]. Zhonghua yu fang yi xue za zhi [Chinese journal of preventive medicine] 54, 1–6 (2020).

5. W. H. Organization, “Non-pharmaceutical public health measures for mitigating the risk and impact of epidemic and pandemic influenza: annex: report of systematic literature reviews,” (World Health Organization, 2019).

6. M. W. Fong, H. Gao, J. Y. Wong, J. Xiao, E. Y. Shiu, S. Ryu, B. J. Cowlings, Nonpharmaceutical Measures for Pandemic Influenza in Nonhealthcare Settings-Social Distancing Measures. Emerging infectious diseases 26, (2020).

7. W. H. O. W. Groups, Nonpharmaceutical interventions for pandemic influenza, international measures. Emerging infectious diseases 12, 81 (2006).

8. F. Matthews Pillemer, R. J. Blendon, A. M. Zaslavsky, B. Y. Lees, Predicting support for non-pharmaceutical interventions during infectious outbreaks: a four region analysis. Disasters 39, 125–145 (2015).

9. Y. Ji, Z. Ma, M. P. Peppelenbosch, Q. Pans, Potential association between COVID-19 mortality and health-care resource availability. The Lancet. Global health 8, e480 (2020).

10. Chinese Clinical Guidance for COVID-19 Pneumonia Diagnoses and Treatment version 6, March 7^th^, 2020. (http://www.nhc.gov.cn/jkj/s3578/202003/d29e176f35ad4b0a80c74c1d347bfbca.shtml).

11. S. Tian, N. Hu, J. Lou, K. Chen, X. Kang, Z. Xiang, H. Chen, D. Wang, N. Liu, D. Lius, Characteristics of COVID-19 infection in Beijing. Journal of Infection, (2020).

12. Report of the WHO-China Joint Mission on Coronavirus Disease 2019 (COVID-19) Pathogen from 2020-02–16 to 2020-02-24.(https://www.cma.org.cn/module/download/downfile.jsp?classid=0&filename=52ef638872004fd9b47104a86dc33fcc.pdf).

13. M. Chinazzi, J. T. Davis, M. Ajelli, C. Gioannini, M. Litvinova, S. Merler, A. Pastore y Piontti, K. Mu, L. Rossi, K. Sun, C. Viboud, X. Xiong, H. Yu, M. E. Halloran, I. M. Longini, A. Vespignanis, The effect of travel restrictions on the spread of the 2019 novel coronavirus (COVID-19) outbreak. Science, eaba9757 (2020).

14. H. Tian, Y. Li, Y. Liu, M. Kraemer, B. Chen, J. Cai, B. Li, B. Xu, Q. Yang, P. Yang, Y. Cui, Y. Song, P. Zheng, Q. Wang, O. Bjornstad, R. Yang, O. Pybus, B. Grenfell, C. Dye, Early evaluation of the Wuhan City travel restrictions in response to the 2019 novel coronavirus outbreak. (2020).

15. B. J. Quilty, S. Clifford, C. n. w. group2, S. Flasche, R. M. Eggos, Effectiveness of airport screening at detecting travellers infected with novel coronavirus (2019-nCoV). Eurosurveillance 25, 2000080 (2020).

16. S. J. Clifford, C. A. B. Pearson, P. Klepac, K. Van Zandvoort, B. J. Quilty, R. M. Eggo, S. Flasches, Interventions targeting air travellers early in the pandemic may delay local outbreaks of SARS-CoV-2. *medRxiv*, 2020.2002.2012.20022426 (2020).

17. Y. Zhang, B. Jiang, J. Yuan, Y. Taos, The impact of social distancing and epicenter lockdown on the COVID-19 epidemic in mainland China: A data-driven SEIQR model study. *medRxiv*, 2020.2003.2004.20031187 (2020).

18. J. Hellewell, S. Abbott, A. Gimma, N. I. Bosse, C. I. Jarvis, T. W. Russell, J. D. Munday, A. J. Kucharski, W. J. Edmunds, F. Suns, Feasibility of controlling COVID-19 outbreaks by isolation of cases and contacts. The Lancet Global Health, (2020).

19. M. Wang, A. Jiang, L. Gong, L. Luo, W. Guo, C. Li, J. Zheng, C. Li, B. Yang, J. Zeng, Y. Chen, K. Zheng, H. Lis, Temperature significant change COVID-19 Transmission in 429 cities. *medRxiv*, 2020.2002.2022.20025791 (2020).

20. M. M. Sajadi, P. Habibzadeh, A. Vintzileos, S. Shokouhi, F. Miralles-Wilhelm, A. Amorosos, Temperature and Latitude Analysis to Predict Potential Spread and Seasonality for COVID-19. *Available at SSRN 3550308*, (2020).

21. Q. Guan, M. Liu, Y. J. Zhuang, Y. Yuan, S. S. Wang, J. Li, Z. Chen, X. L. Yang, Z. R. Tang, H. J. Jia, J. Y. Ma, X. X. Wang, P. G. Tai, J. Li, Y. Zhuang, Y. Hes, [Epidemiological investigation of a family clustering of COVID-19]. Zhonghua liu xing bing xue za zhi = Zhonghua liuxingbingxue zazhi 41, 629–633 (2020).

22. S. Chen, Z. Zhang, J. Yang, J. Wang, X. Zhai, T. Bärnighausen, C. Wangs, Fangcang shelter hospitals: a novel concept for responding to public health emergencies. The Lancet, (2020).

23. K. Mizumoto, K. Kagaya, A. Zarebski, G. Chowells, Estimating the asymptomatic proportion of coronavirus disease 2019 (COVID-19) cases on board the Diamond Princess cruise ship, Yokohama, Japan, 2020. Euro Surveill 25, 2000180 (2020).

24. H. Nishiura, T. Kobayashi, T. Miyama, A. Suzuki, S. Jung, K. Hayashi, R. Kinoshita, Y. Yang, B. Yuan, A. R. Akhmetzhanov, N. M. Lintons, Estimation of the asymptomatic ratio of novel coronavirus infections (COVID-19). *medRxiv*, 2020.2002.2003.20020248 (2020).

25. W.-j. Guan, Z.-y. Ni, Y. Hu, W.-h. Liang, C.-q. Ou, J.-x. He, L. Liu, H. Shan, C.-l. Lei, D. S. Hui, B. Du, L.-j. Li, G. Zeng, K.-Y. Yuen, R.-c. Chen, C.-l. Tang, T. Wang, P.-y. Chen, J. Xiang, S.-y. Li, J.-l. Wang, Z.-j. Liang, Y.-x. Peng, L. Wei, Y. Liu, Y.-h. Hu, P. Peng, J.-m. Wang, J.-y. Liu, Z. Chen, G. Li, Z.-j. Zheng, S.-q. Qiu, J. Luo, C.-j. Ye, S.-y. Zhu, N.-s. Zhongs, Clinical characteristics of 2019 novel coronavirus infection in China. *medRxiv*, 2020.2002.2006.20020974 (2020).

26. S. A. Lauer, K. H. Grantz, Q. Bi, F. K. Jones, Q. Zheng, H. R. Meredith, A. S. Azman, N. G. Reich, J. Lesslers, The Incubation Period of Coronavirus Disease 2019 (COVID-19) From Publicly Reported Confirmed Cases: Estimation and Application. Annals of Internal Medicine, (2020).

27. Z. Hu, C. Song, C. Xu, G. Jin, Y. Chen, X. Xu, H. Ma, W. Chen, Y. Lin, Y. Zheng, J. Wang, Z. Hu, Y. Yi, H. Shens, Clinical characteristics of 24 asymptomatic infections with COVID-19 screened among close contacts in Nanjing, China. Science China Life Sciences, (2020).

28. C. Poirier, W. Luo, M. S. Majumder, D. Liu, K. Mandl, T. Mooring, M. Santillanas, The Role of Environmental Factors on Transmission Rates of the COVID-19 Outbreak: An Initial Assessment in Two Spatial Scales. *Available at SSRN 3552677*, (2020).

29. A. C. Lowen, S. Mubareka, J. Steel, P. Paleses, Influenza virus transmission is dependent on relative humidity and temperature. PLoS Pathog 3, e151 (2007).

30. K. Lin, D. Y.-T. Fong, B. Zhu, J. Karlbergs, Environmental factors on the SARS epidemic: air temperature, passage of time and multiplicative effect of hospital infection. Epidemiology & Infection 134, 223–230 (2006).

